# Longevity of seropositivity and neutralizing titers among SARS-CoV-2 infected individuals after 4 months from baseline: a population-based study in the province of Trento

**DOI:** 10.1101/2020.11.11.20229062

**Authors:** Paola Stefanelli, Antonino Bella, Giorgio Fedele, Stefano Fiore, Serena Pancheri, Eleonora Benedetti, Concetta Fabiani, Pasqualina Leone, Paola Vacca, Ilaria Schiavoni, Arianna Neri, Anna Carannante, Maurizio Simmaco, Iolanda Santino, Maria Grazia Zuccali, Giancarlo Bizzarri, Rosa Magnoni, Pier Paolo Benetollo, Silvio Brusaferro, Giovanni Rezza, Antonio Ferro

## Abstract

**Background:** There are conflicting results about the duration of antibodies induced by SARS-CoV-2, but several studies show a rapid decay in a few months after infection. To evaluate antibody decline, we re-evaluated the presence of anti-SARS-CoV-2 antibodies among individuals found seropositive in a first population survey conducted 4 months before.

**Methods:** All individuals above ten years of age resident in 5 municipalities of the Autonomous Province of Trento, northern Italy, who resulted IgG positive for anti-SARS-CoV-2 nucleocapsid (NC) antibodies in a serosurvey conducted on May 2020 were retested after 4 months. Anti-SARS-CoV-2 antibodies were detected using the Abbott SARS-CoV-2 IgG assay (Abbott Diagnostics, USA) detecting anti-NC antibodies. Samples that gave a negative result were re-tested using the same test plus Liaison SARS-CoV-2 IgG assay (DiaSorin, Italy) to assess anti-spike (S) S1/S2 IgG antibodies. Seroprevalence was calculated as the proportion of positive people on the total number of tested. A neutralizing assay was performed on a subgroup of formerly positives sera using fifty-percent tissue culture infective dose (TCID_50_) as endpoint dilution to produce a cytopathic effect in 50% of inoculated Vero E6 cells culture. In all the analyses a p value < 0.05 were considered statistically significant. Statistical analysis was performed by STATA version 16.1 (STATA Corp., College Station, Texas, USA).

**Findings:** Overall, 1159 out of 1402 initially anti-NC seropositive participants were enrolled in the study. Of them, 480 (41.1%) became seronegative for anti-NC IgG antibodies. When 479 negative sera were tested for anti-S IgG, 373 samples (77.9%) resulted positives. A functional neutralization assay was performed on 106 sera showing high concordance with anti-S antibodies positivity.

**Interpretation:** A decline of anti-NC IgG values was recorded 4 months after the first evaluation. Worth of note, a high proportion of anti-NC seronegative individuals were positive for anti-spike IgG antibodies, which appear to persist longer and to better correlate with neutralization activity.

## Introduction

The presence of neutralizing antibodies induced by natural infection or by an effective vaccine is likely to be predictive of protection. Thus, whether antibody response to the administration of vaccines may confer protection is still undefined, and cases of reinfection by severe acute respiratory syndrome coronavirus-2 (SARS-CoV-2) have been sporadically reported1.

The duration of the immune response after infection is also under investigation. The duration of protection against infection with common human coronaviruses appears to be rather short^2,3^, and there are studies showing declines in IgG antibodies against SARS-CoV-2 among both symptomatic and asymptomatic individuals^4,5^.

Whether memory-B-cell and T-cell responses may still confer protection in individuals experiencing antibody decline to undetectable levels is unknown^6^. In general, controversies exist on the possibility that SARS-CoV-2 infection induces sustained humoral immune responses in convalescent patients following symptomatic COVID-19^7-9^.

The type of antibody response may also play a role. Experimental vaccination against SARS-CoV with NC can induce strong antibody responses that were found to be non-neutralizing^10^. While non-neutralizing antibodies might still exert antiviral activity, for example via the Fc-Fc receptor-based effector function, non-neutralizing NC antibodies may lead to enhanced disease for some vaccine candidates in animal models when neutralizing antibodies are absent^10^. Instead, studies conducted on SARS-CoV-2 and other coronaviruses have shown that the spike protein is the main target for neutralizing antibodies^11-13^.

Thus, assessing the duration of detectable antibody response and changes in the titer of neutralizing antibodies is an important step in order to better understand their dynamics and to predict the duration of protection against SARS-CoV-2 infection.

In order to evaluate the persistence of SARS-CoV-2 antibodies, we repeated a serosurvey in five municipalities of the Autonomous Province (AP) of Trento, Italy, recruiting those individuals who had resulted positive in a large population-based seroprevalence study conducted 4 months before^14^. Moreover, in a subsample of seropositive participants, the antibody neutralizing titre was also evaluated.

## Methods

### Study population and design

As already reported^14^, the study was conducted in 5 municipalities of the AP of Trento, in the northern of Italy, with the highest incidence of COVID-19 confirmed cases.

The Department of Prevention of the Azienda Provinciale per i Servizi Sanitari (APSS) sent a letter of invitation to participate at a second study to all the citizens who resulted to be positive for anti-SARS-CoV-2 antibodies in the serosurvey conducted 4 months before, between May 5 and 15, 2020.

### Serum preparation and storage

Blood samples (5 ml) were collected in Serum Separator Tubes (BD Diagnostic Systems, Franklin Lakes, NJ, USA) and centrifuged at room temperature at 1600 rpm for 10 min. Aliquots were transferred to 2ml polypropylene, screw cap cryotubes (Sorfa, Zhejiang, China) and immediately frozen at −20 °C. Frozen sera were then shipped to the Istituto Superiore di Sanità (ISS) as national reference laboratory for COVID-19, in dry ice following biosafety shipment condition. Upon arrival serum samples were immediately stored at −80 °C.^14^

### SARS-CoV-2 IgG immunoassays for nucleocapsid (NC) and spike (S)

Two commercial CLIA assays, employing either NC or S antigens and designed for high throughput in healthcare settings, were used. In particular, all the serum samples were evaluated by using the Abbott SARS-CoV-2 IgG assay (Abbott Diagnostics, Chicago, IL, USA); sera resulting negative were retested using the DiaSorin Liaison SARS-CoV-2 IgG assay (DiaSorin, Italy). The Abbott Diagnostics anti-NC IgG assay was performed on the Architect i2000SR automated analyser. The analyser automatically calculates SARS-CoV-2 NC IgG antibody concentration expressed as an index value. According to the manufacturer’s instructions, the results were interpreted considering as positive an index of ≥ 1.4 and as negative an index of <1.4.

The DiaSorin SARS-CoV-2 IgG assay is also a two-step CLIA assay for the detection of IgG antibodies against S1/S2 antigens of SARS-CoV-2. The assay was performed on the LIAISON® XL fully automated chemiluminescence analyzer. The analyser automatically calculates SARS-CoV-2 S1/S2 IgG antibody concentrations expressed as arbitrary units (AU/mL). The assay range is up to 400 AU/mL. According to manufacturer’s instructions, values ≥ 15 AU/mL were interpreted as positive, and values ≤ 12 AU/mL as negative; in case of results falling within an equivocal zone in between 12 AU/mL and 15 AU/mL, the test was repeated.

### SARS-CoV-2 neutralizing antibody assay

*In vitro* neutralising activity provides quantitative results as a measure of a functional humoral immune response against SARS-CoV-2. A known amount of SARS-CoV-2 (code 77III, isolated and cultivated at ISS, titre 1×10^5,4^; GISAID accession ID: EPI_ISL_412973) was incubated with different dilutions of the serum sample to determine the dilution at which cytopathic effect on Vero E6 cells (ATCC® CRL-1586) is observed in 50% of infected wells (MN 50%). The detailed protocol is described below: two-fold serial dilutions of serum samples starting at 1:8 dilution up to 1:512 in cell culture medium EMEM (Sigma) supplemented with 1X pen/strep and 2% fetal bovine serum (FBS; Corning) were added to 96-well plates. The mixture of virus (100 TCID_50_) and serum was incubated at 37°C for 1 hour for a total 100 µl. After this incubation period, a solution of 22,000 cells per well in a total volume of 100 µl was added and incubated at 37°C for 5 days.

Finally, the neutralization titer was calculated and expressed as the serum dilution capable of reducing the cytopathic effect to 50% (MN 50%). Positive and negative sera samples and cell culture control together with the virus were added in each test.

### Statistical analysis

The IgG values were summarized by the median and interquartile range (IQR). The differences among IgG values between the first and the second survey were evaluated by the Wilcoxon test. The differences among IgG values between groups (positive versus negative in the second survey) in the first survey were assessed by Mann-Whitney test. The IgG values observed in the first survey were categorised in tree classes: “*weak positive*” (between 1.4 and 3.0), “*medium positive*” (between 3.0 and 5.0), and “*highly positive*” (>5). The McNemar’s test was used to compare frequency on paired data. The concordance between anti-NC, anti-S, and TICD50 was evaluated using the Kappa test (K < 0.20 = “poor”, 0.20–0.40 = “fair”, 0.40–0.60 = “moderate”, 0.60–0.80 = “good”, and 0.80–1.00 = “very good”).

A multivariable logistic regression model was used to determine the relationship between persistent anti-NC IgG in the second serosurvey (positive versus negative) and a set of explanatory variables. The following variables that were significantly associated (p< 0.10) at the univariate analysis were included in the multivariable model: gender, age group, geographical area, presence of symptoms, working in contact with the public and household size, IgG positivity group (weak, medium, high) olfactory and gustatory dysfunctions, fever, weakness, cough, dyspnea, arthralgia, diarrhoea, and abdominal pain and vomit. The likelihood ratio test was used to compare different models.

A subset of anti-NC IgG positive samples was tested with the neutralization test. Assuming a positive proportion of 95% and precision of 4%, 106 samples are required with an alpha error of 5%.

In all the analyses a p-value < 0.05 was considered statistically significant. Statistical analysis was performed by the STATA version 16.1 (STATA Corp., College Station, Texas, USA).

### Ethical approval

Informed consensus for blood collection was obtained from all the participants. The study was approved by the Ethical Committee of the ISS (Prot. PRE BIO CE n.15997, 04.05.2020),

## Results

### Participation in the second survey

Overall, 1159 individuals of the 1402 individuals who resulted seropositive in the first survey (82.7%) were enrolled in the study. All age groups were well represented. The proportion of those who were retested ranged between 72.6% in the age group 20-29 years and 93.1% in the age group 60-69 years.

### Changes in antibody levels against NC

Of the 1159 individuals who resulted initially seropositive 480 (41.4%) seroreverted at the second evaluation. As shown in Figure 1, a statistically significant reduction in the median value was observed in the second survey, from a median of 5.7 (IQR = 3.5) to 1.9 (IQR = 2.8) (p-value < 0.0001 using the non-parametric Wilcoxon signed-rank test).

**Figure 1.**
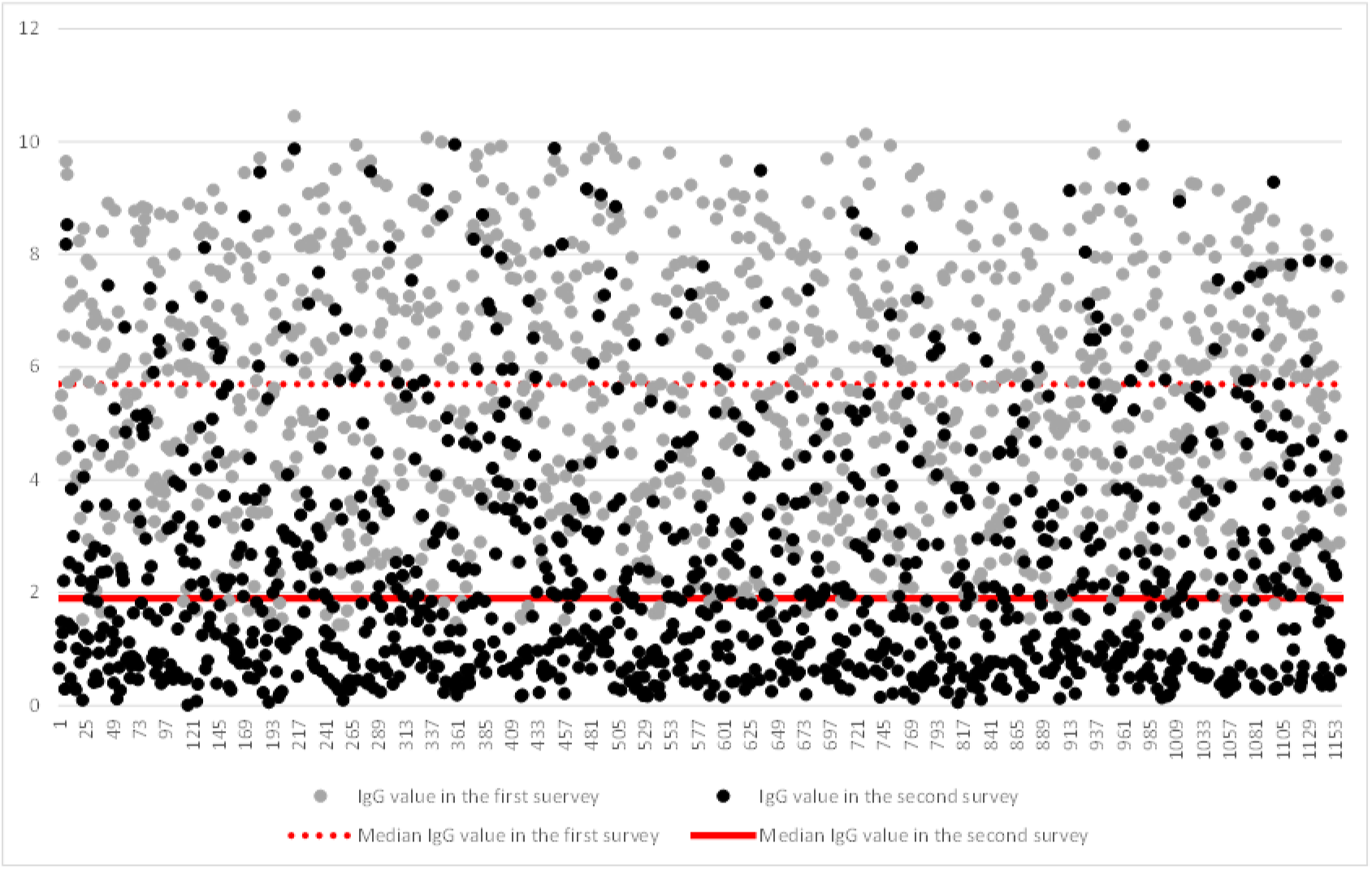
Distribution of the IgG values against SARS-CoV-2 nucleocapsid in the first (grey) and in the second (black) serosurvey.

Comparing the median values in the positive and negative groups, those who seroreverted started from a lower average value (median = 3.6; IQR = 1.9) compared with those who remained positive (median = 7.0; IQR = 2.3) at the second survey; the difference was statistically significant (Mann-Whitney test; p <0.0001).

As shown in Figure 2, when the participants were stratified in three groups in accordance with their anti-NC IgG level at the baseline [i.e., weak positive (with a value between 1.4 and 3), medium positive (between 3 and 5), and high positive (greater than 5)], the median value of the weakly and moderately positive groups decreased below the assay cut-off after 4 months, while the median of the highly positives remained above the cut-off.

**Figure 2.**
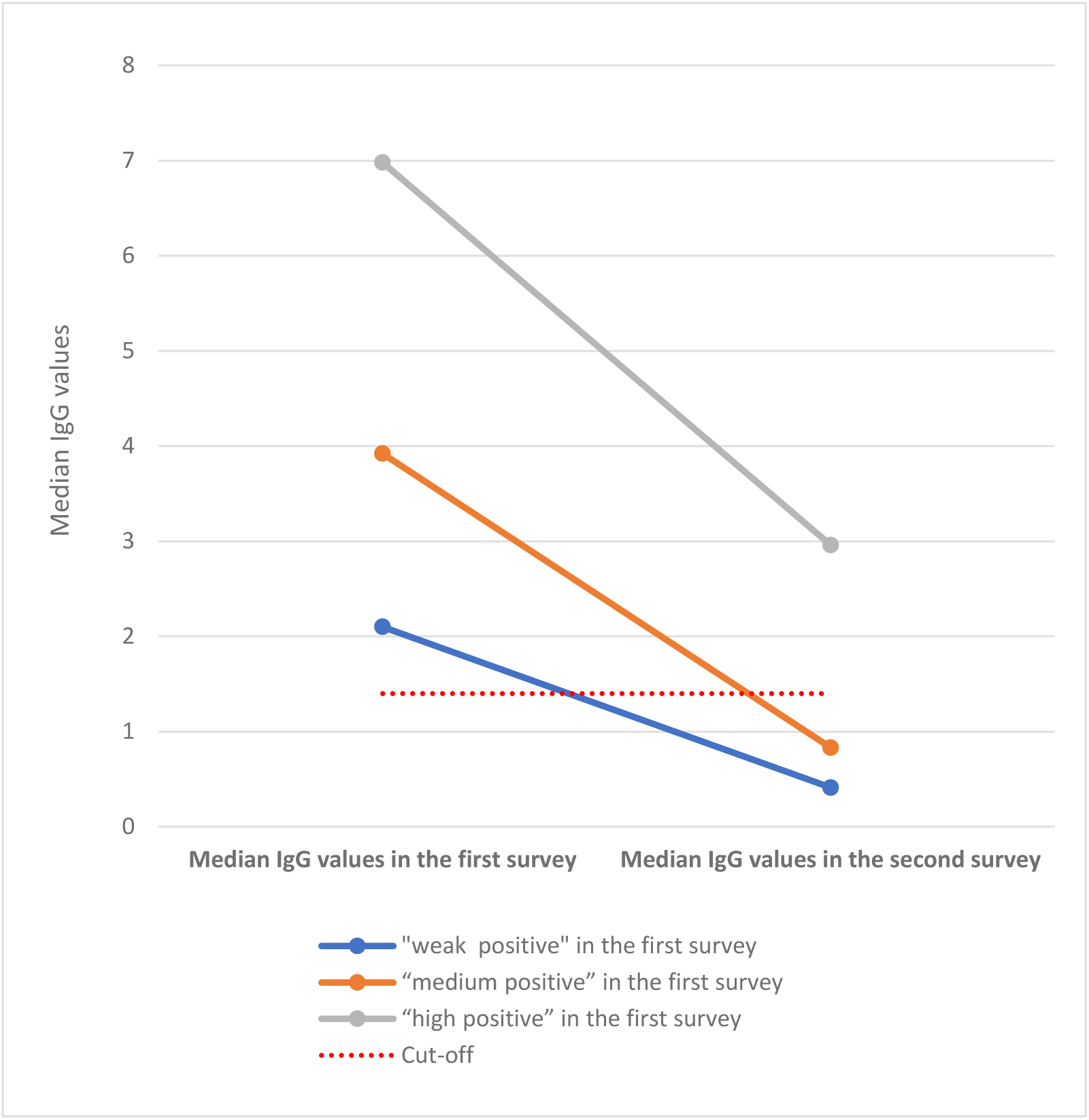
Median of the IgG values against SARS-CoV-2 nucleocapsid in the first and second survey by IgG positivity groups.

### Correlation between anti-SARS-CoV-2 IgG against NC and S proteins and neutralization activity

The samples resulting negative for antibodies against NC in the second study were tested to evaluate the presence of antibodies against the S protein. Since for one sample the available amount of serum was not sufficient for the analysis, 479 available serum samples were tested, and 373 of them (77.9%) resulted positive (Figure 3).

**Figure 3.**
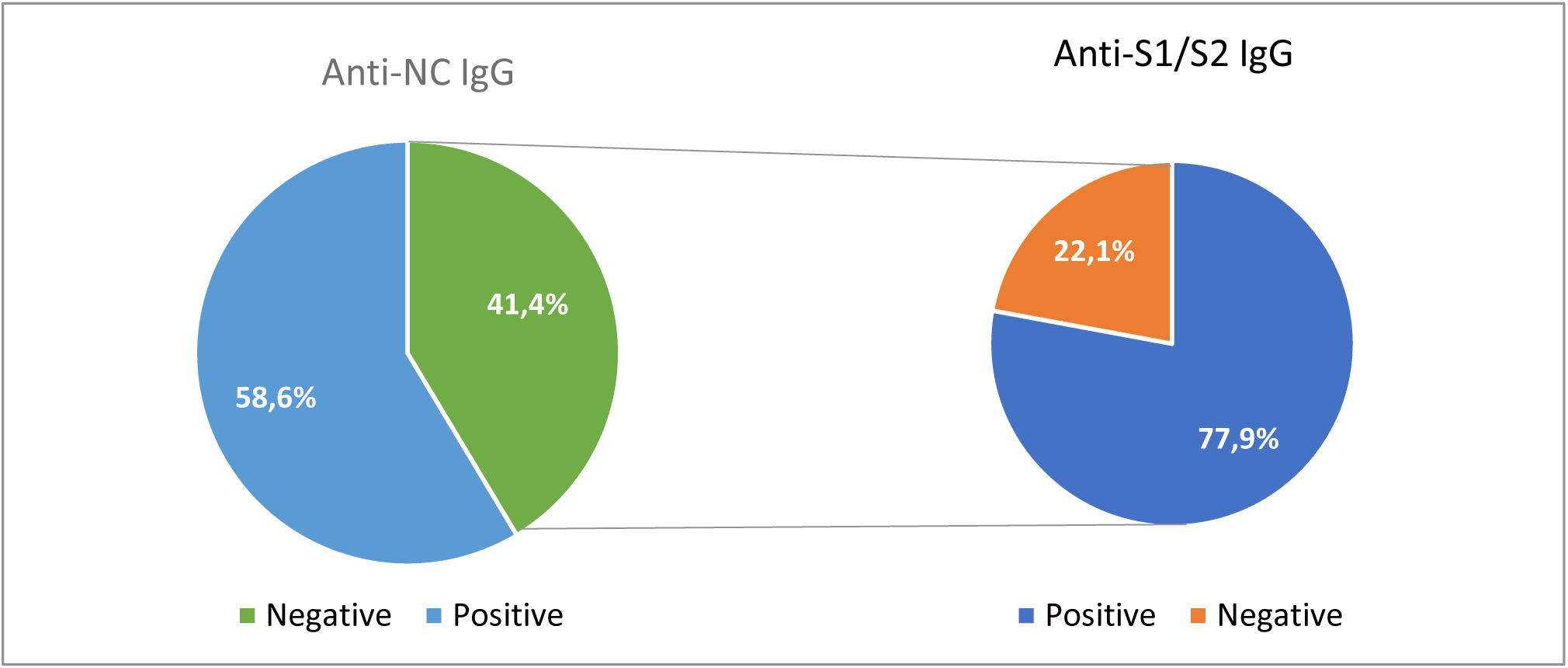
Percentage of anti-spike (S1/S2) IgG antibodies on retested anti-NC IgG negative sera.

### Comparison between serology and functional neutralization assay

A subgroup of 106 sera that were positive for anti-NC-IgG at the first testing were tested for anti-NC IgG, anti-S IgG, and their neutralizing activity 4 months after the baseline. Of the 106 sera, 97 (91.5%) showed neutralizing activity (TCID50 ≥ 1/8), and 9 sera (8.5%) had a TCID50 titer <1/8; 57 (53.8%) were anti-NC positive and 93 (87.7%) were anti-S positive. (Figure 4).

**Figure 4.**
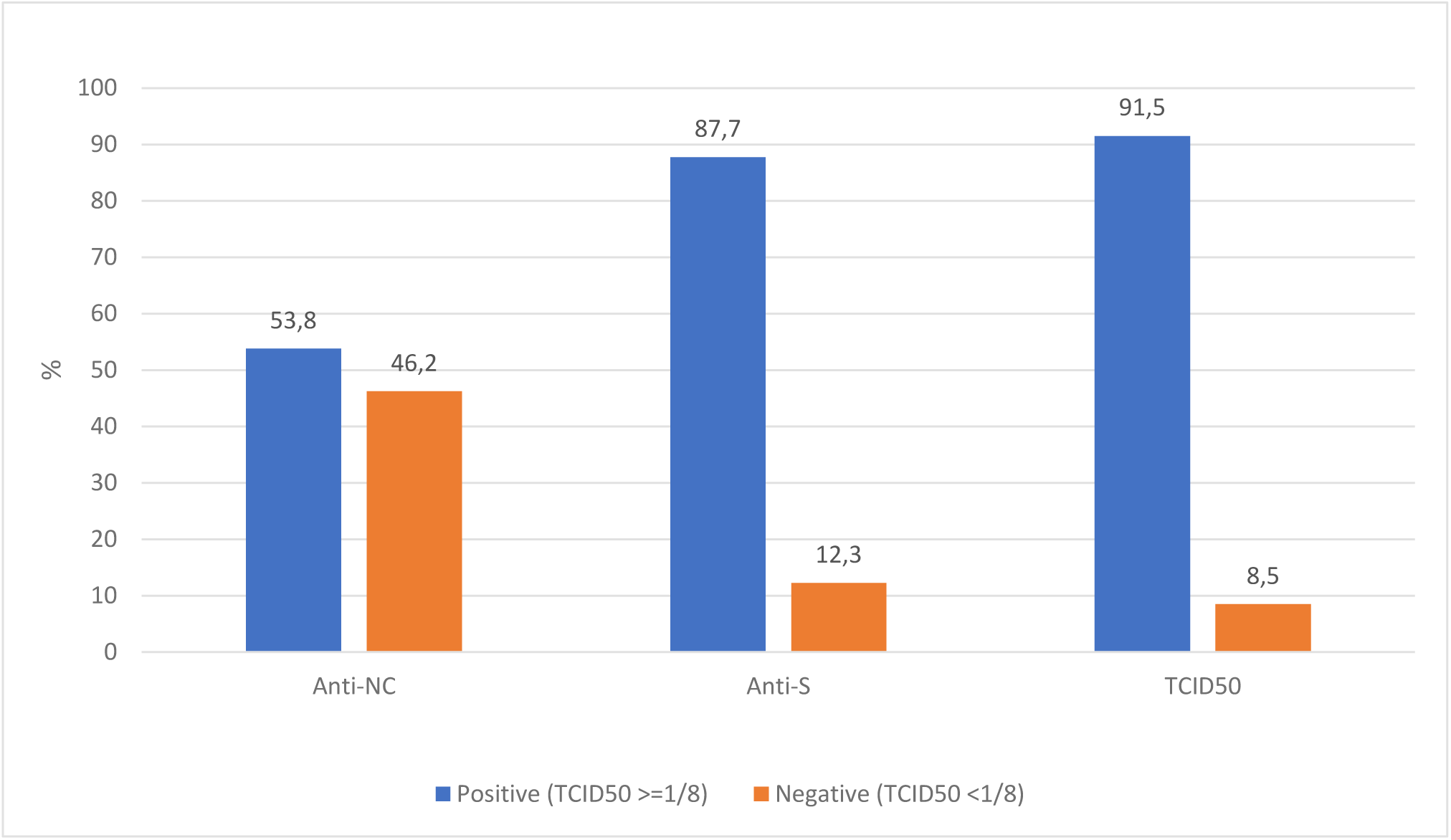
Comparisons between serology and functional neutralization assay.

As shown in Table 1, only 53 sera showing neutralizing activity were anti-NC IgG positive (54.6%) versus 92 (94.8%) which were anti-S IgG positive. Most of the anti-NC IgG negative sera (41out of 49) were anti-S positive (83.7%) and 44 had neutralizing activity (89.8%). Of 93 anti-S positive sera, 92 showed neutralizing activity, confirming a high concordance between anti-S positivity and neutralization activity, as calculated by McNemar’s test.

**Table 1.** Concordance between IgG against NC and S proteins and neutralization activity.

|  | Anti-S + | Anti-S - | p-value* | Agreement | TCID50 ≥ 1/8 | TCID50 <1/8 | p-value* | Agreement |
| --- | --- | --- | --- | --- | --- | --- | --- | --- |
|  |  |  |  | Kappa; p-value |  |  |  | Kappa; p-value |
| Anti-NC + | 52 | 5 | < 0.0001 | 56.6% | 53 | 4 | < 0.0001 | 54.7% |
| Anti-NC - | 41 | 8 |  | K=0.08; p=0.1186 | 44 | 5 |  | K=0.03; p=0.2787 |
| Anti-S + |  |  |  |  | 92 | 1 | 0.1025 | 94.3% |
| Anti-S - |  |  |  |  | 5 | 8 |  | K=0.70; p<0.0001 |

\* McNemar test

High and significative agreement (94,3%) was found between anti-S and TCID_50_ (k=0.70; p<0.0001) (Table 1). To further confirm the concordance, when IgG levels were considered, a good correlation between anti-S and TICD50 was observed (rho-Spearman: 0.84, p < 0.0001) compared with anti-NC/anti-S (rho-Spearman: 0.61, p < 0.0001) and anti-NC/TICD50 (rho-Spearman: 0.56, p < 0.0001).

### Factors associated with persistent anti-NC IgG after 4 months

The multivariable logistic regression model showed that age group, gender, anti-NC IgG level in the first serosurvey, and cough were factors associated with the persistence of anti-NC seropositivity (Table 2). In particular, the individuals with high anti-NC IgG levels in the first serosurvey had the highest probability to be seropositive after four months (OR=69.2). Age above 70 years and cough, as reported during the first survey, were also strongly associated with persistent anti-NC IgG levels.

**Table 2.**
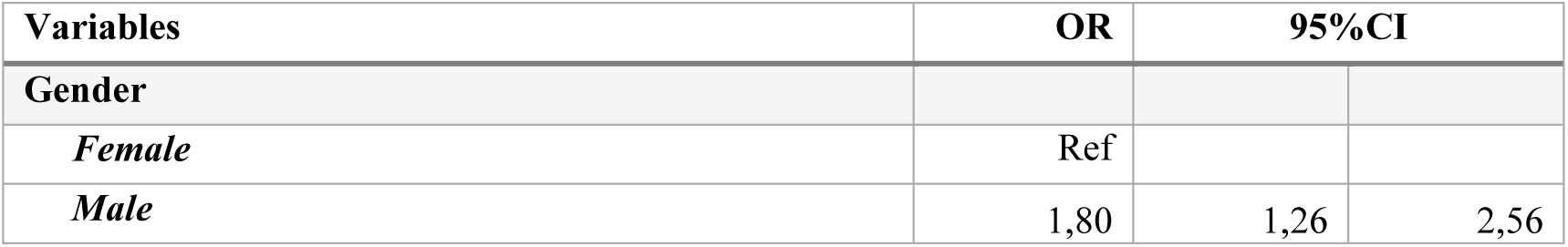
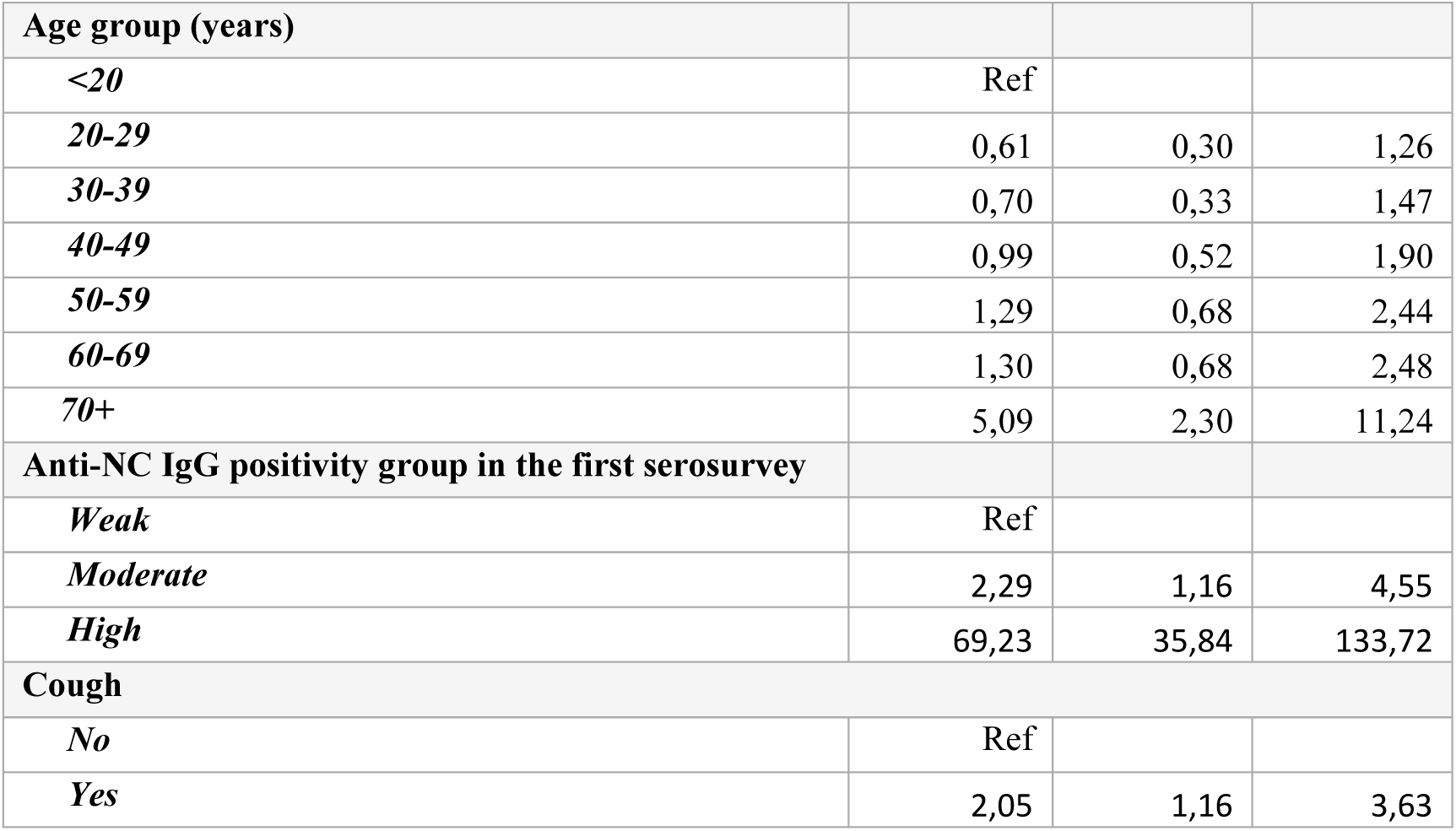
Factors associated with seropositivity (multivariable logistic regression model).

## Discussion

Hereby, we report the results of a repeated serosurvey conducted in five municipalities in the AP of Trento, located in northern Italy^14^. One of the main findings of the second survey, conducted on a large population of initially seropositive individuals, consisted in the rapid decrease of antibodies against the SARS-CoV-2 nucleocapsid. Of the 1159 participants, 41.1% resulted seronegatives by 4 months after the first evaluation. Surprisingly, when we tested the NC-negative serum samples for antibodies directed against the spike protein, we found different results, with most patients still showing seropositivity. To better understand and explain these findings, we evaluated the presence of neutralizing antibodies in a subgroup of previously anti-NC seropositive individuals and found that almost all the sera positives for antibodies against the spike protein were able to neutralize the virus entry into cell lines in vitro.

Correlates of protection have been identified for many viral infections. These correlates are usually based on a specific level of antibodies induced by vaccination or natural infection that significantly reduces the risk of (re-)infection. For some viral infections and vaccines, the kinetic of the antibody response is also known, allowing for a prediction of how long protection will persist^15^.

Previous studies had shown conflicting results. Studies conducted on a smaller number of individuals and/or clinical series reported a decay of neutralizing antibody levels 2 months after infection ^4,5^. These results appear to be consistent with those obtained for other human coronaviruses, such as NL63, 229E, OC43, and HKU1, showing a rapid decay of antibodies directed against the nucleocapsid protein^16^. However, other studies showed different results, with high IgG levels after several months ^7-9,17^. The inconsistency in the results of previous studies could be explained by differences in the study populations (i.e., patients with mild vs moderate or severe disease) or in the use of different methods (i.e., detection of antibodies directed against the nucleocapsid vs whole spike or the receptor binding domain of the spike)^4^. More recently, Seow et al. ^18^ described the longitudinal decline of antibody responses in SARS-CoV-2 infection in sequential sera collected up to 94 days from the onset of symptoms of 65 COVID-19 patients.

In a longitudinal study of RT-PCR confirmed COVID-19 cases, the participants showed a wide range of antibody responses, and a decline in antibodies levels and virus neutralization was observed within three months of the onset of symptoms^18^. For those who developed a low neutralizing antibody response (ID50 100–300) the titers could return to baseline over a relatively short period, whereas those who developed a robust neutralizing antibody response maintained titers >1000 despite the initial decline^18^. The decline of protective antibodies might be explained, to some extent, by the sporadic COVID-19 reinfection that have been reported^19-21^. In a more recent study, detectable neutralizing antibody responses were detected for several months after infection^17^.

To this regard, animal models show that SARS-CoV-2 infection protect from re-infection for at least some time^22,23^. This protection appears to be more pronounced in the lower respiratory tract rather than in the upper respiratory tract^23^.

The types of antibody response may also play a role. Atyeo et al^24^, showed that a predominant humoral response to nucleocapsid protein is associated with poor outcome in patients admitted to hospital, compared to response to spike protein.

Most vaccine candidates elicit responses to S rather than NC protein. Measuring antibodies to S will therefore indicate whether there has been a good response. To this regard, our findings are in good agreement with the study by Wajnberg and colleagues showing that antibody responses to the S protein correlate significantly with SARS-CoV-2 neutralization^17^.

Before drawing conclusions, strengths and limits should be mentioned. Firstly, only 17.3% of individuals did not participate in the survey, thus the refusal rate was low and the possibility of a selection bias was minimized. Secondly, the study was repeated approximately 4 months after the first test; however, a proportion of the participants was apparently asymptomatic and others reported having had symptoms suggestive of COVID-19 sometime before the survey. Thus, the 4 months represent the minimum interval of time elapsed between the virtual date of infection and the second test. Thirdly, although the serological assay we used is assumed to have high sensitivity and specificity, the occurrence of some false positive or false negative results influencing the reliability and consistency of the results could not be completely ruled out.

In conclusion, we found a general antibody decay over time, with a relatively high proportion of initially SARS-CoV-2 seropositive individuals losing their anti-nucleocapsid antibodies by 4 months after the first positive test. However, most of these individuals still had neutralizing anti-spike IgG antibodies, suggesting a potential long-term duration of protective immune response. This finding may have important implications in the choice of the target for antibodies persistence over the time together with the potential effectiveness and long-term protection of immune responses induced by vaccines and on herd immunity. Further studies are needed to understand whether persistence of anti-spike, potentially neutralizing antibodies, is actual correlates of long-term protection.

## Data Availability

NA

## Acknowledgments

The authors would like to thank the study’s participants.

The authors thank the Serosurvey Study Group for COVID-19 in the AP of Trento: Mattevi Elisabetta, Dalla Valle Lorenza, Endrizzi Luca, Pecoraro Lucia, Varesco Andrea, Brida Elisa, Daldoss Alessia, Sebis Claudia, Bruno Zanon, Mauro Bandera, Laboratory Dept., APSSTrento; Riccardo Flavia and Patrizio Pezzotti, Dept. Infectious Diseases, Istituto Superiore di Sanità; Pocher Massimo, Sannicolò Renzo, Manica Andrea, Fogarolli Angela E-healthcare Solution Service, APSS Trento;Maroni Veronica, Chizzola Elisa Privacy Office, APSS Trento; Sforzin Simona Primary Care Director APSS Trento, and the entire staff of primary care involved in the study.

## Funds

APSS sustained the expenses for the IgG assays on collected sera.

## Authors contribution

P.S., A.B. together with A.F. were responsible for the conception and design of the study; G.F. and P.S. coordinated the analysis on sera; P.L., P. V., A. N., A. C., I.S., M. S., I. S., S. F., E. B., C.F. performed the analysis on sera; S.P. M.G.Z., G.B., R.M., P.P.B., organized the samples and data collection; A.B. performed the statistical analysis; S. B. and G. R. help in the discussion of data; G.R. revised critically the manuscript; P.S. wrote the manuscript. All the authors revised and approved the manuscript.

